# COVID-19: Comparison between 8-days and extended 4weeks outbreak periods through socioeconomic and natural factors

**DOI:** 10.1101/2020.06.11.20107086

**Authors:** Sana Ullah, Jianghua Zheng, Zhengkang Zuo, Feizhou Zhang, Ke Shang, Wenjie Yu, Yu Fu, Chuqiao Han, Yi Lin, Kaiwen Jiang, Shanlin Sun, Yiyuan Sun, Shoujiang Zhao, Lei Yan

## Abstract

Since mid-March 2020, global COVID-19 pandemic has experienced an exponential growth in process from sporadic to sudden outbreaks. This paper selects the 8-day surge data of daily cases, death and recovery rates (March 19-26, 2020) from 18 countries with severe pandemic situation to discuss the impact of 9 factors of both socioeconomic and natural on the pathogen outbreak. Moreover, the paper also elaborates analysis and comparison of relatively slow 4-week (February 1-29, 2020) data of China’s surge cases to determine the relationship between social and natural factors and on the spread of pandemic, which provides an effective reference for delaying and controlling the pandemic development.

## Introduction

Since March, the COVID-19 pandemic is spreading exponentially. Although many countries have adopted response strategies, the pandemic has so far spread in more than 200 countries/territories/areas around the world, which infected more than one million people and caused tens of thousands of deaths [1].

The study of relevant factors on the sudden spread of pandemic has become the most direct means to effectively delay and control the outbreak. From the recently published literature, the first focus is on the impact of social factors, especially government control on the pandemic situation of COVID-19[2-18]; According to the data of pandemic situation, a model is developed to predict the effect of government intervention measures to slow down the pandemic situation and then how long these active intervention measures need to be maintained to control the situation [19-20]. When social factors such as government intervention have an effect, the impact of natural factors to slow down the pandemic and the degree of impact is begun to be concerned. Only few scholars have considered the impact on the COVID-19 pandemic from the perspective of natural factors, especially meteorological conditions, but there is no unified answer due to different experimental conditions [21-25]. At the same time in China, due to strict social interventions, the whole process from a small amount to an outbreak was greatly delayed to 4 weeks in February 2020, which can be a special reference in delaying the surge of cases. Based on this, this paper selects the number of daily average cases from different countries across the globe for during the 8-day surge period and compare it with the 4-week data of similar surge period in China to quantitatively evaluate the relationship between socioeconomic and natural factors with pandemic spread. Trying to reveal, social, natural or both factors play a major role in the global massive transmission and in guiding control of COVID-19, and further how to deal with it.

## Materials and methods

### Data collection and study plan

The study assessed the real cause of the Novel Covid-19 spread both in China and across the globe. Moreover, the assessment plan is designed to consider both environmental and social factors and to draw their nexus with the spread out. Apart from that, some considerations how to retain the virus spread from further spreading are also taken into considerations. For experimental purpose, 30 cities of the China and 18 countries across the globe are selected. Those countries and Chinese cities are selected in such a way to draw the relationship between environmental factors, i.e. temperature, humidity, aerosol level and vegetation, and social factors, i.e. population density, per capita income, and people to people contact. Besides, data from some countries out of the selected pool are also used for the model validation. Moreover, worldwide daily reported cases are also collected which are used for geostatistical analysis.

As for as the data collection is concerned, various data sources are used to gather the relevant data used in the assessment of relevant study, as shown in Table S1. Besides, a retrospective population data is collected from the online data repositories for the 2019 Novel Corona virus Visual Dashboard operated by the Johns Hopkins University Center for Systems Science and Engineering (JHU CSSE; https://github.com/CSSEGISandData/COVID-19), and worldometer https://www.worldometers.info/coronavirus/fcountries. These online systems are providing real-time coverage of the COVID-19 outbreak all over the world including different states in China. The databases have obtained data from numbers of sources mentioned in their website. These are publicly available online data without an identification of infected patients directly obtained from public health authorities or by state media or WHO reports. We collected Chinese statistics concluding daily cases, daily death rate, daily recovery rate from DX Doctor COVID-19 Global Pandemic Real-time Report https://ncov.dxy.cn/ whose data sources are WHO, CDC and local media reports.

### Data compilation

As the outbreak of the COVID-19 occurred around December 2019 and the first confirmed case had been identified in January 2020, and in this regards the Chinese authorities — about that unknown pneumonia detected in Wuhan, China—for the first time reported to WHO (World Health Organization) on the 31^st^ December, 2019. Adding to this, WHO declared the outbreak as Public Health Emergency of International Concerns on 30^th^January[26]. While keeping in view the prevailing situation around China and worldwide, the data is compiled in sections. Firstly, the collection of data is mainly focused on two different time periods, i.e. from Feb 09-29, 2020 and March 19-26, 2020 for both inside China and for the selected 14 countries, e.g. USA, Italy, Germany, France, Spain, Iran, Bahrain, Kuwait, Thailand, Malaysia, Singapore, South Korea, Japan, and Australia. The reason for selection of two different time periods is that during February the outbreak was on its peak around China but its impact was minimal in rest of other countries, while in March the case has been reversed. Moreover, during this period, due to well management of Chinese authorities and the development of healthcare system the spread has control to a large extent, and no such new cases has been recently reported. On top of that, globally the number of cases is increasing with each passing day. Secondly, to validate the disease spread model, the daily reported cases of countries, i.e. USA, Germany, Italy, Spain, UK, France, Switzerland, Belgium, and Netherlands from the date of first case till March 26 are collected. Finally, to draw the geographical picture of the COVID-19 pandemic, worldwide reported cases on a monthly bases, i.e. from virus outbreak till the end of March are also compiled.

As for China, daily cases, daily death, daily recovery data of 23 provinces and 4 municipalities from January 22 to March 31 were collected. The pandemic parameters of each capital cities are estimated based on Equation *(1-3)* for the lack of accurate data in each city. Assuming value “a” as the number of cumulative cases of one provincial capital city while “b” as that of corresponding province until now, then the time-series Daily Cases of each city from January 22 to March 31 can be estimated by multiplying p(case) to the time-series daily cases of corresponding province. The calculated dataset is shown in Table S5.

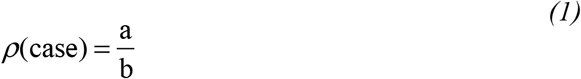

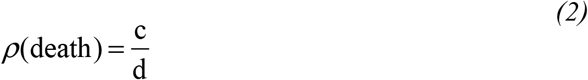

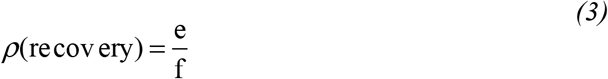

### Statistical analysis

In order to identify the relationship between selected factors, i.e. both social and natural factors with daily recorded infected cases, daily recorded deaths, and daily recorded recovery cases, a descriptive analysis was performed. In this regard, relationship among the selected factors and variables are drawn via regression analysis. Moreover, due to the lack of data for the spread of COVID-19 with respect to people to people contacts, a statistical virus infectious model is developed to show how the virus spread among the people with respect to time. For this purpose, a relationship is drawn between people contacts with respect to time and then taking into consideration the COVID-19 mortality rate from an infected patient to healthy people. On top of that, model is validated via comparing model results —of the multiplication rate of people contacts with respect to time —with the real time COVID-19 spread data for the selected countries as discussed in the previous section. Besides, that some scenarios are assumed for reducing people to people contact and finally its impact on the virus spreading rate.

### Assessment of social contacts with respect to COVID-19

As the COVID-19 virus is very rapidly spreading across the globe in no time. With every single day, the number of virus infected patients is increasing, and the challenges for the governments to cope with it, are increasing too. Since its eruption from Wuhan China, and elsewhere in the world, public health experts are trying to gauge the potential of the COVID-19 by calculating the pathogens basic reproduction number. Since its eruption from Wuhan China, and elsewhere in the world, public health experts are trying to gauge the potential of the covid19 by calculating the pathogens basic reproduction number. In this regard, the introduced term RO 1.5 and 3.5 by the MRC Center for Global Infectious Disease Analysis at the Imperial College London, which pronounced as “R naught” is an estimate of the infection rate that how many healthy people one contagious person will infect. As discussed earlier, from the MRC Center for Global Infectious Disease Analysis at the Imperial College London, the virus RO rate of 1.5 means that if an average of four people is infected with the COVID-19 virus, they would spread the virus among 6 more people, and they further would spread among 9 more people. On top of that, with RO rate of 3.5, those 4 people on average would infect 14 more people followed by 49 more people and so on so forth. Until the infection is contained or run its course, the disease will exponentially will increase [27]. So, the virus reproduction rate with respect to time is of great importance and needs careful considerations. At this point, if it is assumed that one infected person is in contact with a healthy person and after getting infected healthy person those further get infected other health people which is a continues process and make a chain of series in time domain. Keeping in view of that, a virus infectious model is developed, with an assumption to assess the virus reproductivity with time and how to minimize the reproduction rate.

For simplicity, a sample case of population density of 512-person km^-2^ is taken as sample data. It is assumed that population is uniformly distributed over the geographical area, and each person on the geographical plane occupy the center of its occupied area. The distance between two neighboring persons on the geographical plane is the Euclidean distance between them, as shown through following formula:

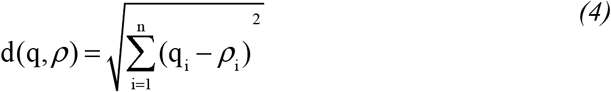

It is further assumed that a normal healthy person can cover an average distance of 8km hr^-1^. Further, in the virus infectious model, it is assumed that each person can only move either in forward or sideways directions, and at each time step can only approach to a single person. Through some basic mathematical calculations, it calculated that an average distance between two neighboring people is 44.19 m, whereas the average distance covered @ of 8km hr^-1^ becomes 133m min^-1^. So, it is also assumed that prior to simulating the model, only a single person is COVID-19 contagious patient. All people will be in travelling at the same time step, in this way only a single person will come in contact to other. Each person will be in contact with other for some predefined timing, e.g. initially for about 5 minutes, which means that after approaching the second person, the first person will spend 5 minutes with him then they will see off each other, and will heading towards the next geographical locations for another contact person and so on so forth. Initially, the modeling period is set for about 70 minutes, which means that people on their will can freely meet with each other during the stipulated time by following the set criteria as discussed before. At the end of the modeling time, following data is achieved which presented in the (Table S2). After successfully developing the virus infectious model, it is further analyzed for the virus reproductive rate.

### Development of scenarios for minimizing social contacts

For such purpose, with the help of virus infectious model, different scenarios are taken into consideration for minimizing the people to people contact either to increasing the travelling time between two predefine people or to confine them at a place for longer period of time as per contact. In first scenario, people to people traveling time is increased from 30sec to 2 minutes and then to 20 minutes (Table S2), which means to create some hurdles against people moments and people to people contacts shouldn’t be as easy as it is during the normal social contacts, whereas in the second scenario, some predefine restrictions are imposed on the people moments, i.e. 10, 20, 50 and 75 % (Table S3)to asses it impact on the people contacts.

## Results and discussion

### Effect of socioeconomic factors at very-short explosion period in coping COVID-19

Amongst the socioeconomic factors, population density and per capita income are tested with those selected variables from global countries. For the spread of virus outbreak through human contact, geography and population density may be the influencing factors for the transmission and, indirectly to human mortality [28], i.e. 1918-19 influenza pandemic which was one of the worst pandemics in the history with an estimated global mortality between 20 to 100 million [29-30]. While analyzing the spread of COVID-19 pandemic with respect to population density, it is found that there is no such significant relationship with the number of daily cases, death and recovery rates (Fig. 1-a, b, c). Among those selected countries, the population density of Singapore and Bahrain are higher, but the number of cases being reported from these countries is lower than those with lower population densities. The reason may be because of strict government policies for reducing the people movements, which is especially taken in Singapore—the country to be approach either by airplanes or ships which can be the source of entering of virus into the country, e.g. the entry of influenza virus in India during the 1918-19 influenza pandemic through the port of Bombay on the west coast [30]. In this regard, Singapore has greatly restricted the entrance of foreigners into the country. On top of that, the Singapore government has also announced “Satellite City” policy to reduce population pressure to maximize the localization, and avoiding long-distance movement and congestion due to shopping, medical treatment, schooling, etc. This policy also reduces the spreading of virus among people to a large extent [31].

**Fig. 1.**
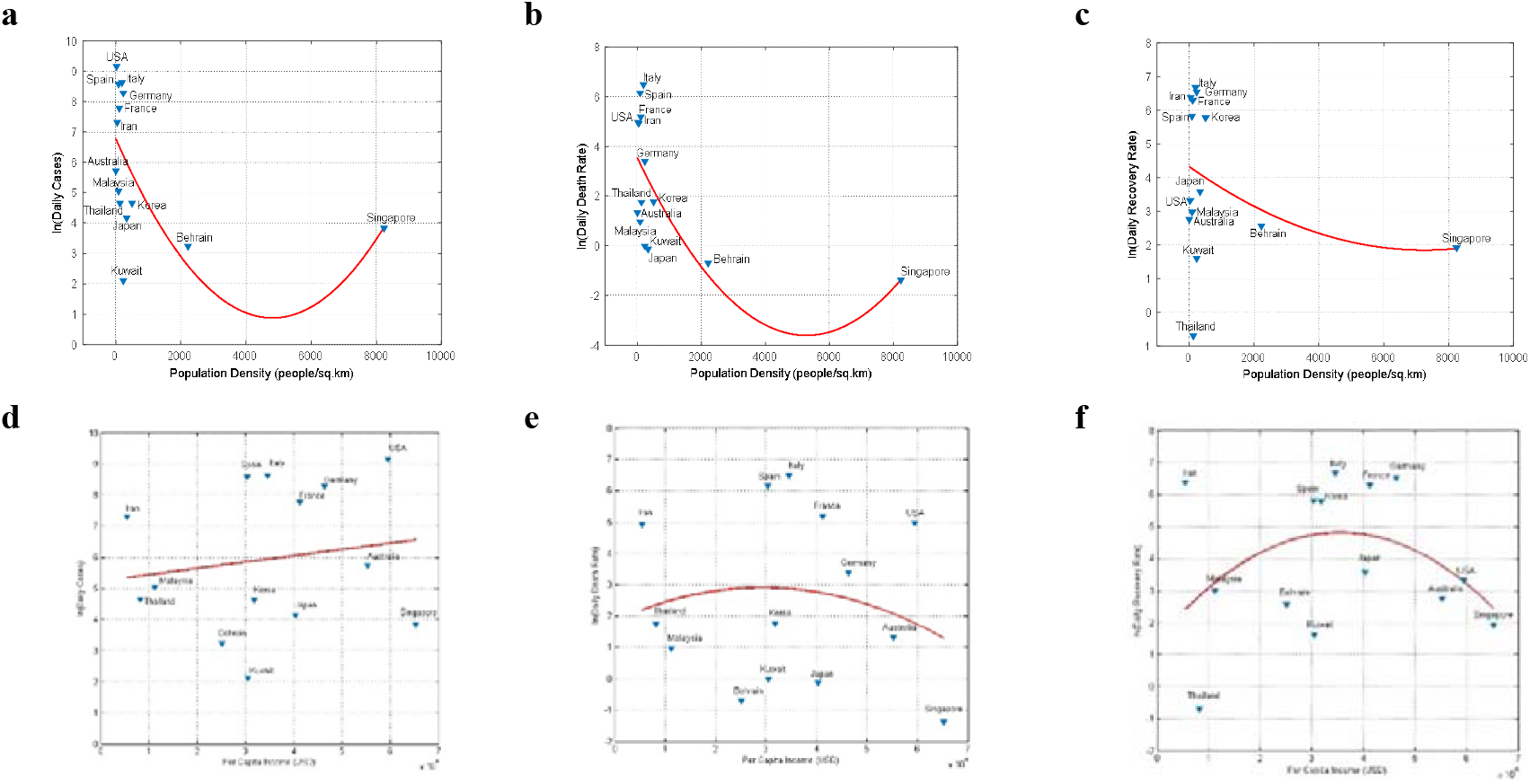
**Effect of Socio-economic factors on the COVID-19 parameters**, i.e. relationship between (**a, b, c**) population density, (**d, e, f**) per capita income vs. daily cases, death rate and recovery rate, respectively. Correlation accuracy evaluation is listed in Table S5.

Next, we tried to build the relationship between the economic conditions with virus spread. Obviously, economics can play a pivotal role in the burden of infectious diseases. Compared to poor countries, rich countries can spend more capital on immunizations, control of disease vectors, and treatments following infections, which allow people to live for longer period and having healthier lives. As we are living in a complex ecological world where we live with animals and other creatures, and nearly two-third of pathogens and parasites that infect humans involve interactions with animals as vectors or alternative hosts [32]. These include some of the worst chronic diseases, i.e. malaria, cholera, plague, etc. In this aspect, for the first time, the pathogen of COVID-19 casing infections in humans also spread from bats. Based on the above facts, and to assess the immunity level against the COVID-19 amongst the selected countries, we compared its impact between economically poor and rich countries, and to draw the relationship through comparing per capita income (USD) with number of daily cases, deaths and recoveries from infection. The results show (Fig. 1-b) that data points are very scattered, indicating number of daily cases has weak relationship with income rate. Even rich counties with better health facilities are more prone to get infected from the COVID-19. In the same way, the relationship between income rate with daily deaths and recovery (Fig. 2-e, f) is also very weak. The same pattern is observed here as in Fig. 1-b. As a whole, the correlation between the pandemic parameters and per capital income is weak.

**Fig. 2.**
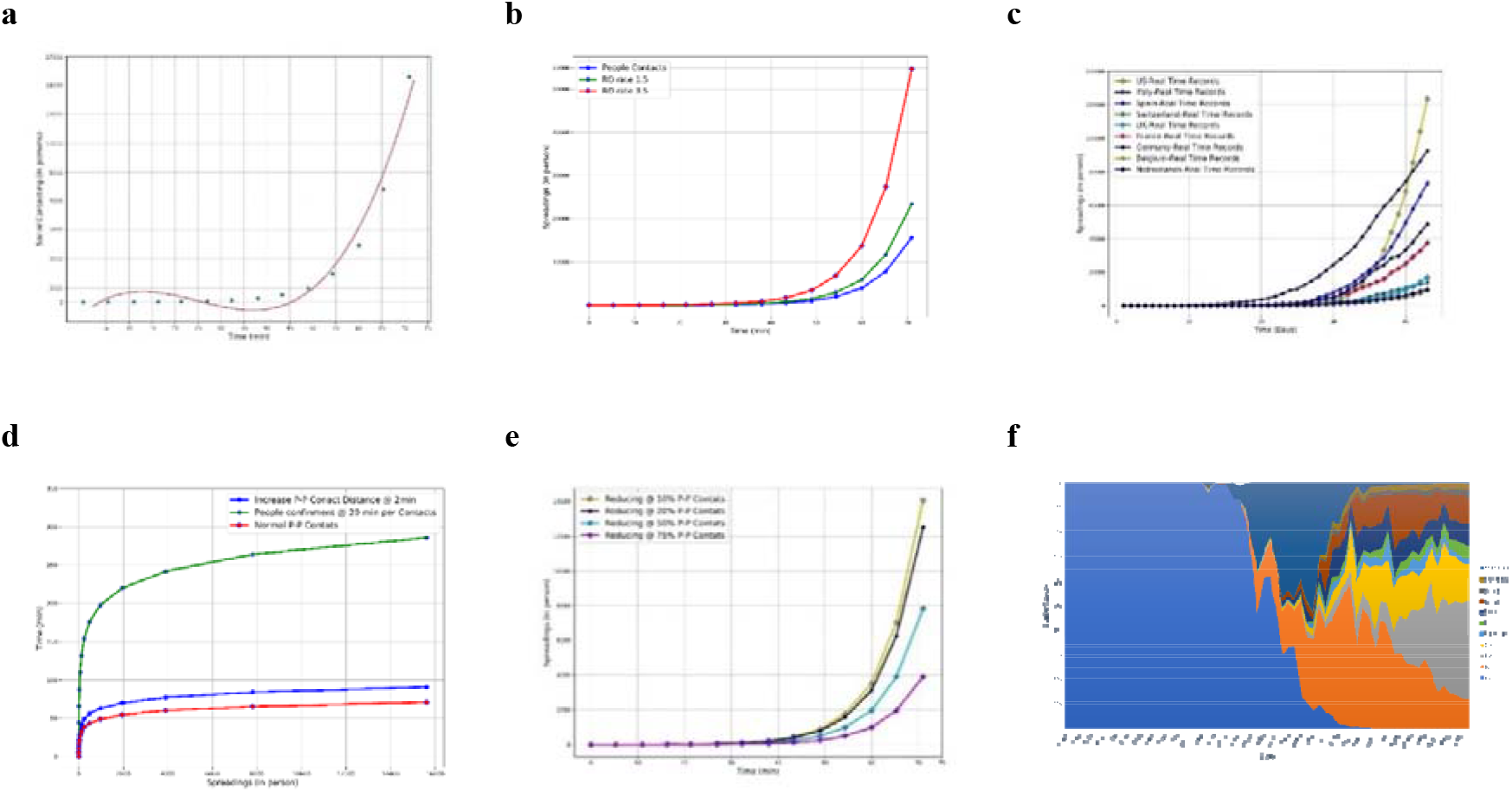
Assessment of virus infection rate through people to people contact model **(a)** number of people contacts in time domain, **(b)** relationship between RO 1.5 & 3.5 with people contacts per unit time, **(c)** plotting real time data for model validation, **(d)** delaying people to people contacts @ 2 & 20 minutes, **(e)** reducing people contacts for about 10, 20, 50, 75%, respectively **(f**) countrywide daily status of the reported cases from reporting

As for as the transmission route of infectious diseases are concerned, social contact pattern among individuals is a key ingredient in the realistic characterization and modeling of pandemics [33-35]. Since social networks are typically seen as a medium for the spread of disease and disease risk factors, but social relationships can also reduce the chance of chronic and potential infectious diseases [36]. On top of that, modeling of infectious diseases transmitted by the respirator or close-contact route such as, pandemic influenza is increasingly used to determine the impact of possible interventions [37].Quantification of human interactions relevant to the disease transmission, i.e. virus spread through social contacts is central to predict disease dynamics [38]. Yet the data, in this regard, for quantifying the social contact patterns relevant to the worldwide spread of COVID-19 available. Keeping in view, we developed a simple modeling approach to draw people to people contacts and then the virus reproduction rate—with some predefined rates, i.e. RO 1.5 and 3.5 (Table S3)— among those contacted people in time domain. From graphical results (Fig. 2-a), it shows an exponential behavior in the time domain. The time is divided in to four quarters. The graph behavior during the first two quarters is statically increasing, while in the third quarter the graph trend drastically changes which indicates in the increasingly replication of people contacts. Onward, considering the people to people contact model as virus infectious model (Fig. 2-b), both RO rates are smoothly increasing with the number of people contacts with time, but at the end of third quarter of the stipulated time, the RO rates started to replicate rapidly and the gap between all three lines is so obvious. That means, the third quarter is very crucial stage for starting the multiplication of the virus spreading within the community on a massive scale. Further, this graphical output is compared with the real time data collected from different countries for the validation procedure. From comparison between the modeling results and real time data (Fig. 2-c), it is obvious that virus spread is showing more or less the same pattern with respect to time. The time to multiplication on a massive rate, from real time data, starts in the end of third quarter of time for all countries except China, South Korea, and Iran, where the virus multiplied massively even before that period—third quarter of time. The reason for outbreak on such a massive scale in China and South Korea at such an early stage is due to the extensive people moment on the eve of New Year (lunar calendar) across the country. As for the Iran is concerned, the massive spread at such early stage is due to large gatherings of people for religious actives across the country, i.e. the Kum city which became the second epicenter after Wuhan China.

So far, the fact is established that virus’s multiplication on a large scale starts during the third quarter of time, but here the most import concern is that how to reduce severity of the outbreak and to normalize the graph trend on a smooth path rather than rapid inclination. For such purpose, with the help of virus infectious model, different scenarios are taken into consideration for minimizing the people to people contact either to increasing the travelling time between the contact of two predefine people or to confine them at a place for a longer period of time as per contact.

**Scenario 1:** In the first scenario, increasing the people to people traveling time from 30 sec (the normal set time for modeling purpose) to 2 minutes and then to 20 minutes (Fig. 2-d), it shows that from start till the 50^th^ minute, the number of people to people contacts are the same, onward that exponential multiplication per unit time starts. On top of that, it is quite obvious that the delay in people contacts from 30 sec to 2 minutes and from 2 to 20 minutes, the multiplication rate delays for about 10 and 150 minutes (in the beginning), and about 20 and 200 minutes (end), respectively. Which shows that with no restrictions on the people movements and social gatherings, virus spreading is exponentially increase in no time, whereas during the restrictions it takes more time to reach the same level of severity. The delay in timing will reduce pressure on the medical care facilities being available for people treatment, and infected people can be treated with some ease. This method can greatly be applied to reduce the people physical contacts which eventually will have the impact on the number of viruses infected people.

**Scenario 2:** In the second scenario, it is observed that restriction imposition reduces people to people contacts to a large extant. From Fig. 2-e, during normal conditions, people to people contacts multiplication starts onward 50^th^ minute at rate of about 1000 people per contact, whereas during 10, 20, 50 and 75% restrictions it is recorded as 880, 781, 488 and 244 people contacts, respectively. Moreover, the reduction in people contacts increasing with respect to time, obvious gap between lines of the graph can be observed. The maximum number of reductions at the end of simulation observed for 75, 50, 20, 10 and 0 % reduction in people contacts are about 3900, 12493, 14056, 7808, 15600 people contacts (Table S4), respectively. Beyond this condition is the complete lockdown of masses, i.e. 100% restrictions mean complete ban on people to people contacts and travelling is minimized to zero level. Moreover, this condition has been seen in many cities of China including the Wuhan City (Fig. S2) soon after the outbreak of COVID-19, and China successful controlled over the situation. The outcome of those complete restriction in China with respect to no restriction or partial restrictions in various countries is shown in the (Fig. 2-f).

### Effect of natural factors at very-short explosion period in coping COVID-19

Apart from socioeconomic factors, we also analyzed the relationship between natural factors, i.e. temperature; relative humidity, aerosol and vegetation cover with those selected variables globally.

As for the impact of temperature is concerned, it shows (Fig. 3-a, e, i) significant negative correlation between daily cases, death and recovery rates, respectively. Moreover, temperature during the selected period amongst the selected countries varies between 5 to 35 ° C, and on contrary, countries with temperature above 15 °C having is no severe pandemic situation, indicating lower temperatures are more conducive for the survival and transmission of virus, which is consistent with the findings of Sajadi [39]. With the increase in temperatures in the northern hemisphere and with the implementation of control measures, the spread of the pandemic is expected to be controlled to a large extent. In contrast, in the southern hemisphere, more care needs to be taken in further spread of pandemic [40].

**Fig. 3.**
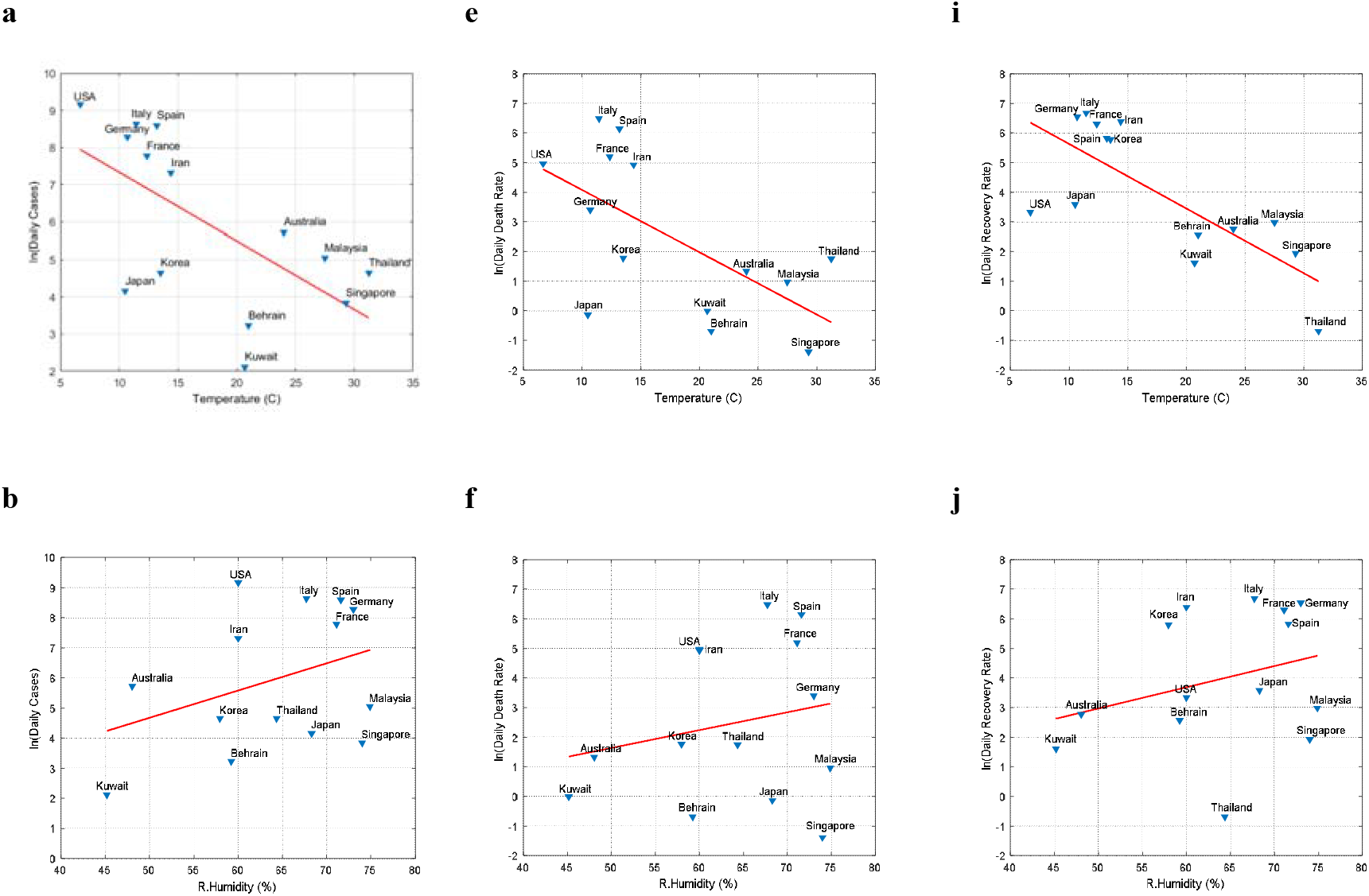
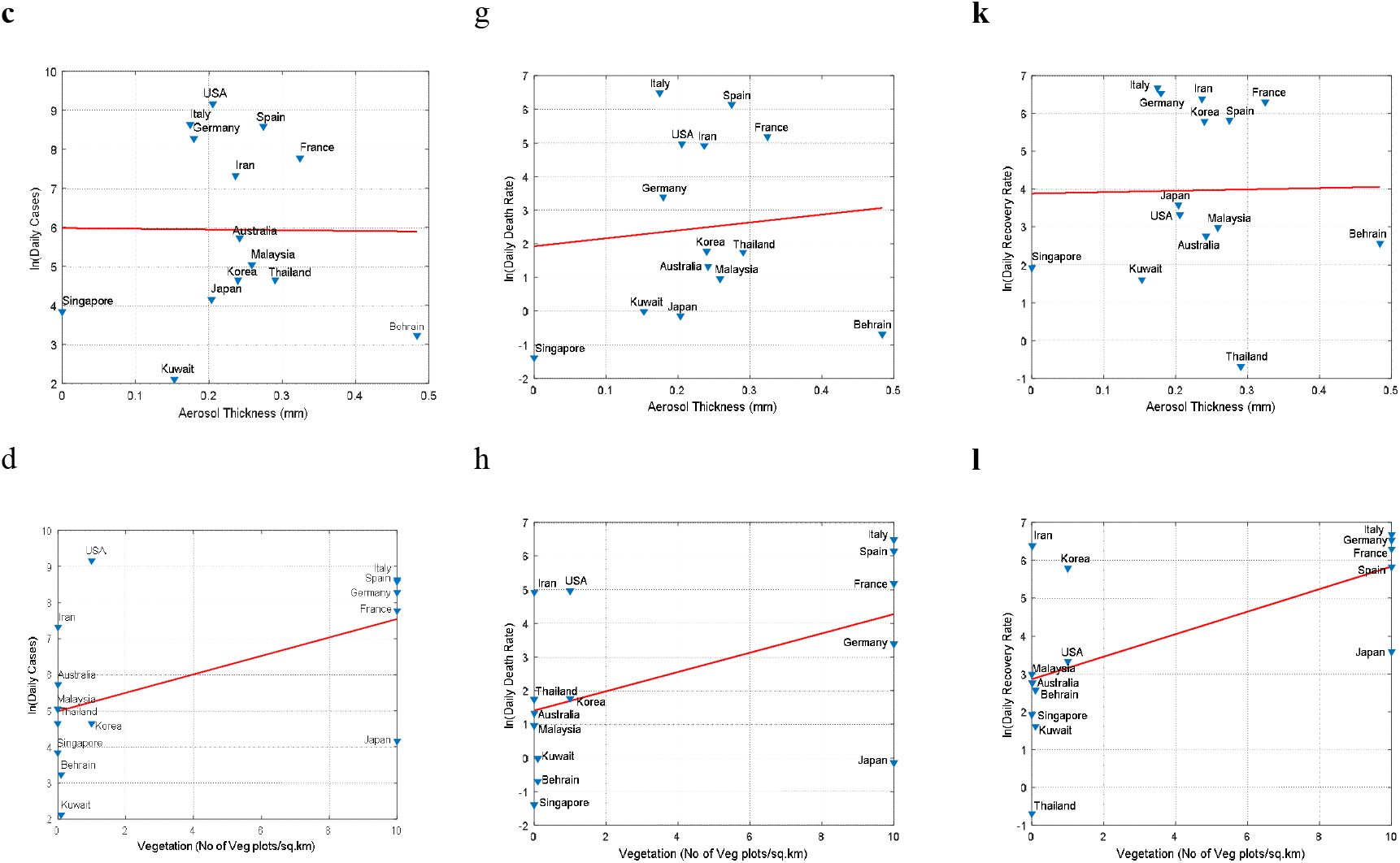
**Effect of Natural factors on the selected COVID-19 parameters**, i.e. relationship between **(a, e, i)** temperature, **(b, f, j)** relative humidity, **(c, g, k)** aerosol, **(d, h, l)** vegetation vs. daily cases, death rate and recovery rate, respectively. Correlation accuracy evaluation is listed in Table S5.

Researches confirmed that respiratory infection increases during unusually cold and humid conditions [41]. Graphically, result (Fig.3-b) indicates no-significant relationship between daily cases and increasing humidity. Despite of having same relative humidity, USA, Iran, South Korea and Bahrain showing varied number of daily cases. Also, Thailand, Japan, Singapore, Malaysia (Asia), and Italy, Spain, Germany and France (Europe) with same relative humidity having different number of daily cases. In addition, similar trend (Fig. 3-f, j) between relative humidity with death and recovery rates are observed. Therefore, low humidity can play important role in suppressing the infection, but it doesn’t necessarily mean that countries with high humidity have severe outbreaks.

Some people show symptoms of COVID-19 without any direct contact in COVID-19 infective which some scholars concluded for inseparable nature of virus spread from aerosol [42]. However, Fig. 3-c shows weak correlation between aerosol and daily cases. Singapore with low aerosol level (approaching 0 mm) shows the higher number of daily cases as compared to Bahrain (about 0.48 mm). Besides, Australia along-with most of the Asian countries except Iran shows low daily cases as compared to European countries including USA. In addition, similar relationships (Fig. 3-g, k) between aerosol and death and recovery rates are observed.

On the part of vegetation cover, it shows non-significant relationships those comparing variables (Fig. 3-d, h, l). Except Japan, countries with higher vegetation cover showing higher number of cases, death and recover rates, and also with low to moderate vegetation cover, countries show varying response to those variables.

### China’s longer explosion period in coping COVID19

The COVID-19 explosion period comprises 8-days as compared to China of 4-weeks. The pandemic growth in China is showing a linear trend as compared to globally. In contrast to global situation, the surge period hit China in February, whereas the situation is normalized to much extent in March. For comparison between surge periods in China and globally, February data is considered. From data, it shows that per capita income & population density have positive correlation with daily cases (Fig.4-a, d), negative & no correlation with death rate (Fig.4-b, e), and weak correlation with recovery rate (Fig.4-c, f), respectively. Moreover, about 80% and 90% cities with low per capita income (<10000 USD) and low population density (< 1000 people/km^2^) showing better situation, i.e. average daily cases < 2 (Fig.4-a, d), respectively. So, high per capita income doesn’t have any advantageous reflections. More gathering and less isolation offset the advantage of high medical level. Lack of awareness about isolation is attributed to culture and customs, but at present it is effectively adopted to minimize the pandemic and to reduce pressure on existing medical conditions. Besides, big cities with high population density had high mobility before enforcement of lockdown, especially on the eve of Spring Festival (Chinese New Year). In contrast, about 57% and 70 % cities with per capita income (<10000 &> 10000 USD) showing the average death rate of >0.05 and < 0.05% % (Fig.4-b), respectively. Similarly, about 67% and 60% cities with population density (<1000 &>1000 people/km^2^) showing the average death rate of < 0.05 % (Fig.4-e), respectively. Lastly, each half cities with per capita income (<10000 &>10000 USD) showing average recovery rate > 3.1% (Fig.4-c), respectively. Similarly, 55% and 70% cities with less population density (<1000 &>1000 people/km^2^) showing average recovery rate > 3.1 and < 3.1% (Fig.4-f), respectively.

**Fig. 4.**
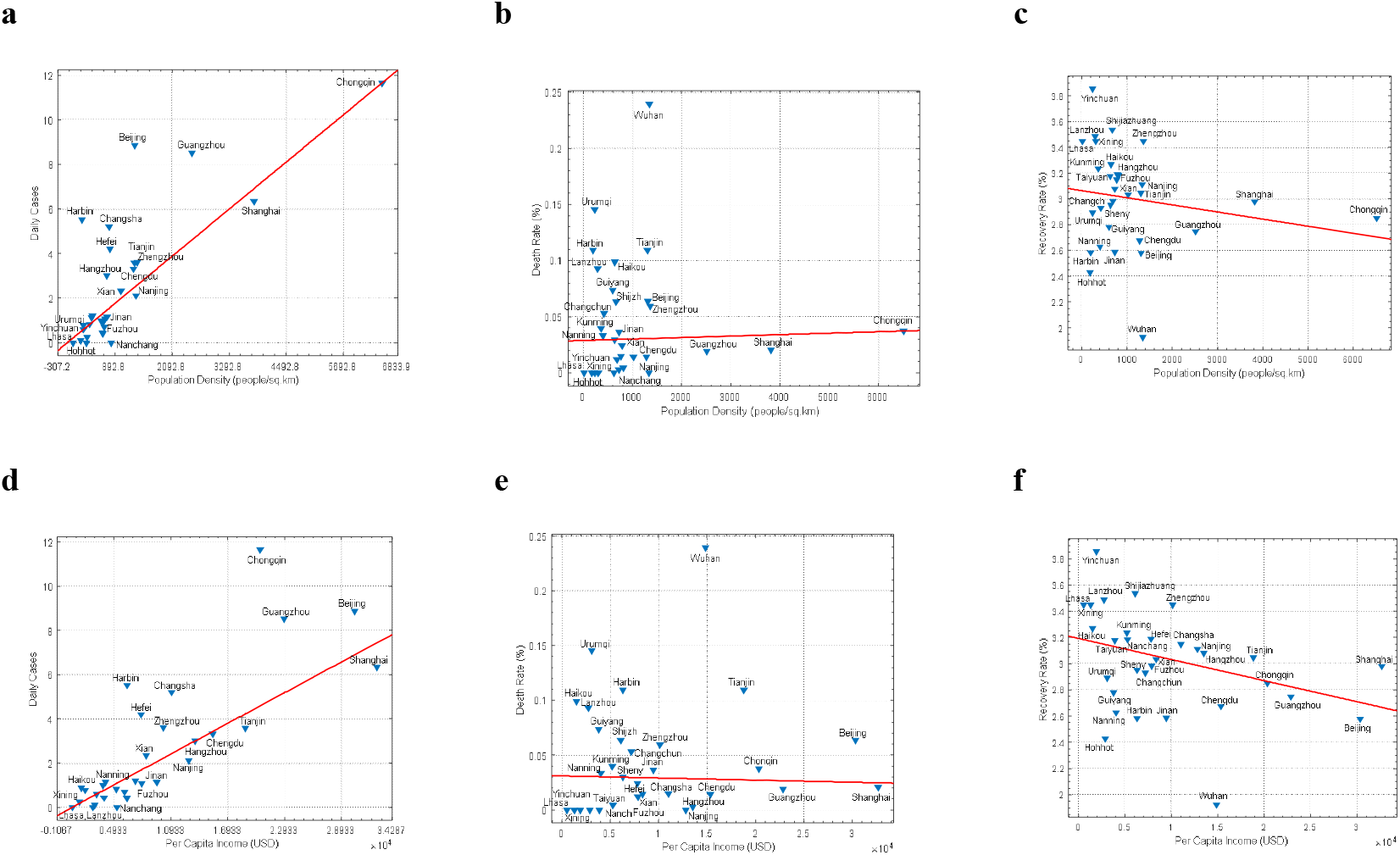
**Effect of Socio-economic factors on the COVID-19 parameters**, i.e. relationship between **(a, b, c)** population density, (**d, e, f**) per capita income vs. daily cases, death rate and recovery rate, respectively.

In addition, to compare the nexus between natural factors with those variables, it shows (Fig. 5-a) positive correlation with daily cases. The temperature of the selected cities during the selected period varies between −13 to 22 °C, but most of cities concentrated in between 1-15 °C. For the daily cases, about 56 % cities reporting less than 2 cases whereas the remaining 46 % are between 2 to 12 cases per day. Therefore, the temperature effect on Corona virus cannot be ignored. Moreover, on the part of humidity, it shows strong positive correlation with the daily cases, but the number of daily cases with respect to humidity is not uniform, e.g. cities which are having the same relative humidity (such as 78%) conditions, having the varied spread rate. On the whole, China’s increased pandemic amongst the selected cities is mainly concentrated with relative humidity of 57% to 78% (Fig.5-b).

However, under the same conditions, there are some cities where the pandemic situation is growing slowly, so other natural factors also have to be considered, i.e. aerosol and vegetation cover. In February, the most severely affected citers are concentrated with the aerosol optical thickness of 0.2 to 0.4 mm (Fig.5-c), but there are no distinct relationships indicating that the virus could spread by aerosol. Moreover, the vegetation cover of a city affects the local temperature, which directly or indirectly affects the spread rate of the virus. The growth trend of the pandemic is mainly distributed between 1 to 30% & over 90% (Fig.5-d). Too high or too low vegetation coverage maybe inhibits the growth rate of the pandemic.

**Fig. 5.**
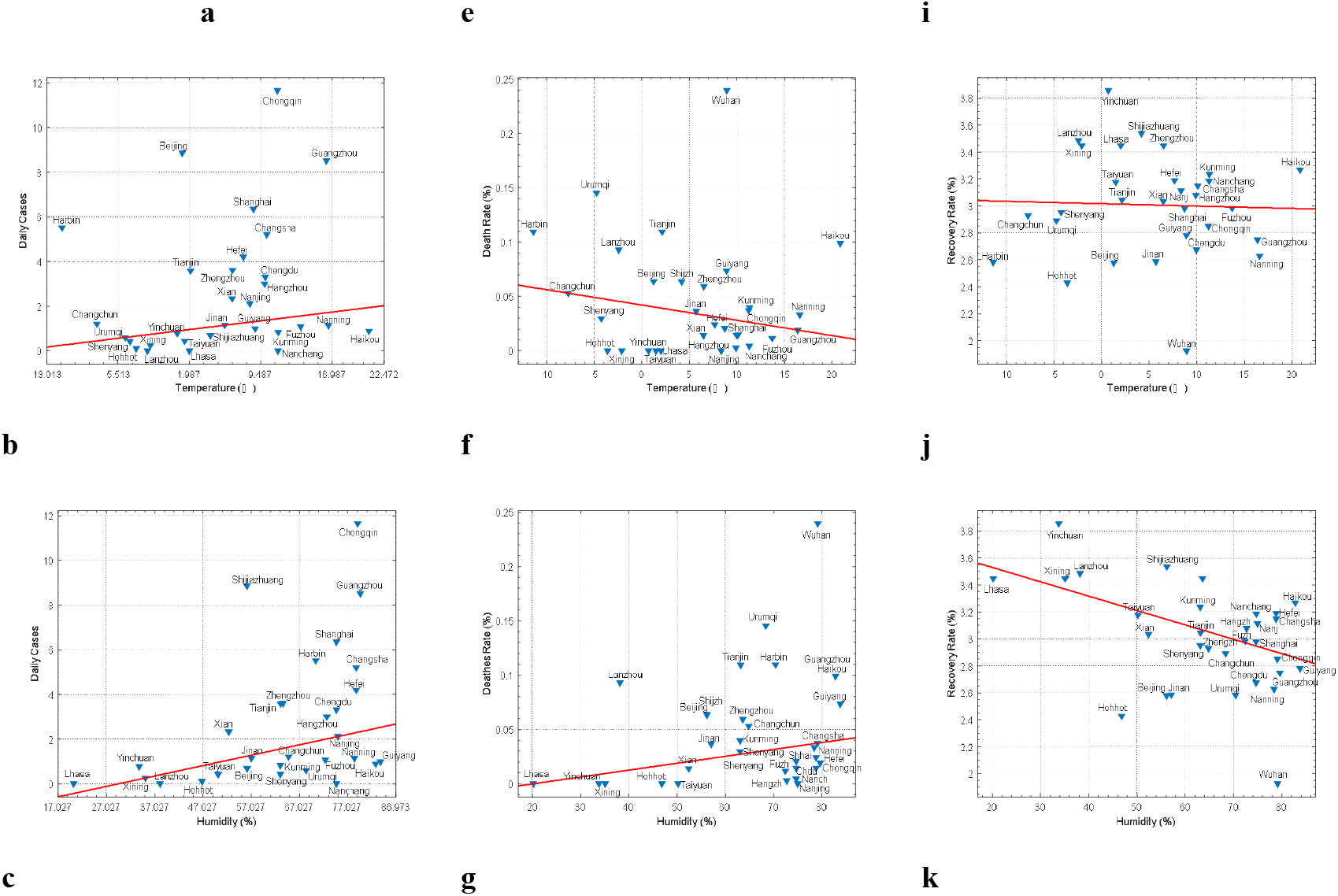
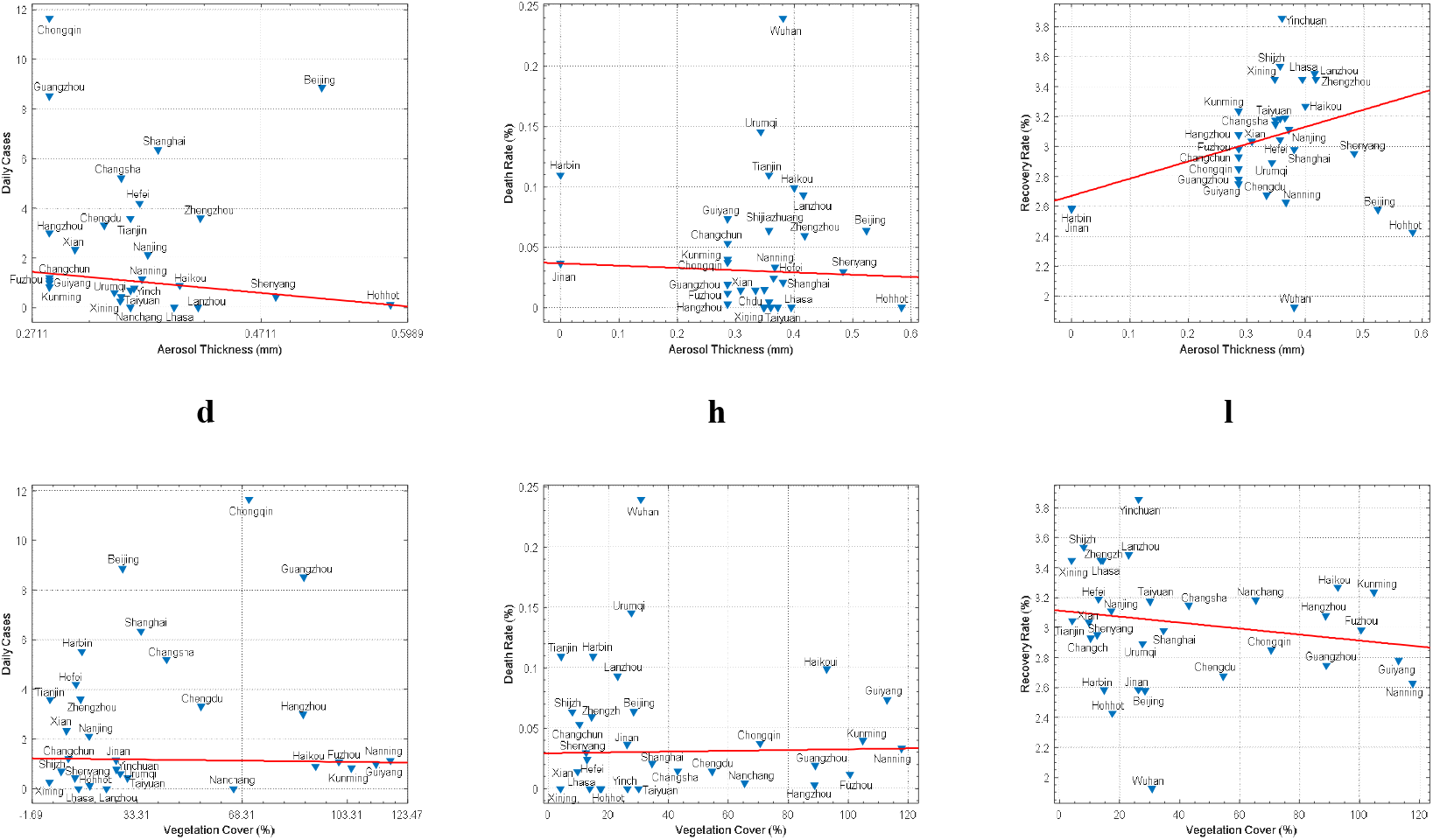
**Effect of Natural factors on the selected COVID-19 parameters**, i.e. relationship between (a, e, i) temperature, (b, f, j) relative humidity, (c, g, k) aerosol, (d, h, l) vegetation vs. daily cases, death rate and recovery rate, respectively. Correlation accuracy evaluation is listed in Table S6.

Moreover, temperature shows relatively negative correlation with death rate, and the temperature of cities with high death rate (> 0.05%) is mainly concentrated between 1-10 ^C^(Fig.2-e). However, the death rate keeps a positive correlation with humidity, and cities having high death rate (> 0.05%) are mainly concentrated between 57% to 78% relative humidity conditions (Fig.2-f). The aerosol optical thickness of cities with high death rate (> 0.05%) is mainly concentrated between 0.3 to 0.4 mm (Fig.2-g). The vegetation cover of cities with high death rate is mainly concentrated between 0-30 (Fig.2-h). There is non-significant correlation between recovery rate and natural factors (Fig.2-i, j, k, l).

### Wuhan as special case in coping COVID-19

Apart from analyzing countrywide spatial distribution, in depth assessment of the Wuhan also took place Fig. S2. The average daily cases reported from January 22 to March 31 are 723, with 501 prior to government control (January), 1524 during the surge period (February) and 22 in March. To slow down the pace of COVID-19 across the China is mainly attributed to large scale vaccination, testing, quarantine, lockdowns, stay-at-home policy and wearing masks for all people. However, our research findings show the temperature nexus with the development of outbreak. Generally, the increase in temperature in Wuhan shows inverse relationships with daily cases between February (2-10 □) and March (14 □ and above) (Fig. S2a). Though temperature in not the main contributing factor to the pandemic, on the contrary, it will slow down the pace of rapid spread. However, the humid environment may help the pandemic spread. For example, during the surge period with average relative humidity (75% −90%) (Fig.S2b), Wuhan even reported low cases days, indicating that virus spread resulted to combined environmental factors. Also, during the surge period, visibility (0-10 km) shows the same impact as that of temperature, and with exceeding 12 km reported lesser number of cases (Fig. S2c). Finally, the atmospheric pressure exceeding 1020 hPa has exacerbated the virus spread, and about 92% of days in March had atmospheric pressure below 1020hpa (Fig. S2d). Therefore, the data of Wuhan (Jan. 22 to Mar. 31) shows high temperature, dry and good air quality, and low atmospheric pressure may helped to slow down the pandemic spread.

As whole, our findings reveal that selected socioeconomic and natural factors are less influencing on the spread of COVID-19 pandemic. Although, comparing with global, these factors show stronger correlations (**R^2^)** with selected pandemic variables in China (Table 1). But still strict government interventions of social distancing and isolation policy in China has delayed the surge period for about 4-weeks as compared to 8-days global surge, and which could serve as example the world in coping COVID-19 successfully.

**Tab 1.**
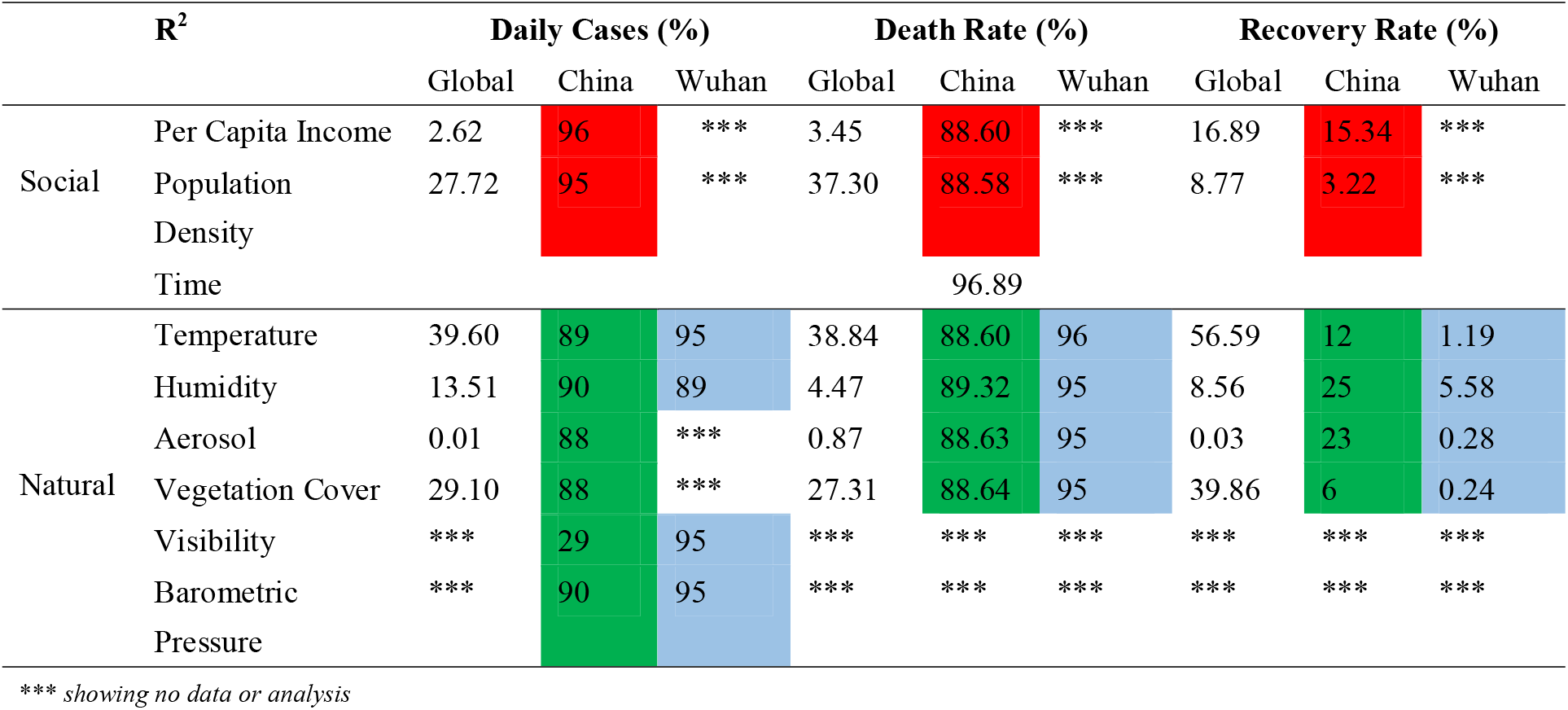
Comparison of the correlation between pandemic variables with socioeconomic and natural factors for Global, Chinese cities, and Wuhan, respectively.

## Data Availability

All the data used in the manuscript can be found in the supplementary materials.

https://github.com/CSSEGISandData/COVID-19

https://www.worldometers.info/coronavirus/#countries

https://ncov.dxy.cn/

https://www.who.int/

https://worldpopulationreview.com/countries/countries-by-density/

https://www.ceicdata.com/en/countries

https://gis.ncdc.noaa.gov/maps/ncei/summaries/daily

https://weatherspark.com/

https://disc.gsfc.nasa.gov/information/glossary?keywords=giovanni%20measurements&title=Giovanni%20Measurement%20Definitions:%20Aerosol%20Index

https://www.givd.info

## Acknowledgments

We really thank to anonymous free online data providers, i.e. Johns Hopkins University Center for Systems Science and Engineering online data repositories (COVID-19), Weather Spark, National Oceanic and Atmospheric Administration (NOAA), National Aeronautics and Scientific Administration (NASA) Earth Science Data, China national and local health and construction commission daily information release (COVID-19), China statistical yearbook, etc., which helped this study to complete successfully in very short time.

## Author contributions

S. U.: conceptualization, formal analysis, methodology, visualization, and writing; J.Z.: project administration; Z.Z.: formal analysis, methodology, visualization, and writing; F. Z.: methodology, writing; K.S.: formal analysis, visualization, and writing; W.Y.: methodology, and writing; F.Y.: formatting; C.H.: data downloading; Y.L.: data downloading; K.J.: data downloading; S.S.: data compilation; Y.S.: data compilation; S.Z.: data compilation; L.Y.: supervision, fund acquisition, and resources.

## Data and materials availability

All data are available in the main text or the supplementary materials.

